# Masks Do No More Than Prevent Transmission: Theory and Data Undermine the Variolation Hypothesis

**DOI:** 10.1101/2022.06.28.22277028

**Authors:** Katia Koelle, Jack Lin, Huisheng Zhu, Rustom Antia, Anice C. Lowen, Daniel Weissman

## Abstract

**Background:** Masking serves an important role in reducing the transmission of respiratory viruses, including SARS-CoV-2. During the COVID-19 pandemic, several perspective and review articles have also argued that masking reduces the risk of developing severe disease by reducing the inoculum dose received by the contact. This hypothesis – known as the ‘variolation hypothesis’ – has gained considerable traction since its development.

**Methods:** To assess the plausibility of this hypothesis, we develop a quantitative framework for understanding the relationship between (i) inoculum dose and the risk of infection and (ii) inoculum dose and the risk of developing severe disease. We parameterize the mathematical models underlying this framework with parameters relevant for SARS-CoV-2 to quantify these relationships empirically and to gauge the range of inoculum doses in natural infections. We then identify and analyze relevant experimental studies of SARS-CoV-2 to ascertain the extent of empirical support for the proposed framework.

**Results:** Mathematical models, when simulated under parameter values appropriate for SARS-CoV-2, indicate that the risk of infection and the risk of developing severe disease both increase with an increase in inoculum dose. However, the risk of infection increases from low to almost certain infection at low inoculum doses (with <1000 initially infected cells). In contrast, the risk of developing severe disease is only sensitive to dose at very high inoculum levels, above 10^6^ initially infected cells. By drawing on studies that have estimated transmission bottleneck sizes of SARS-CoV-2, we find that inoculum doses are low in natural SARS-CoV-2 infections. As such, reductions in inoculum dose through masking or greater social distancing are expected to reduce the risk of infection but not the risk of developing severe disease conditional on infection. Our review of existing experimental studies support this finding.

**Conclusions:** We find that masking and other measures such as distancing that act to reduce inoculum doses in natural infections are highly unlikely to impact the contact’s risk of developing severe disease conditional on infection. However, in support of existing empirical studies, we find that masking and other mitigation measures that reduce inoculum dose are expected to reduce the risk of infection with SARS-CoV-2. Our findings therefore undermine the plausibility of the variolation hypothesis, underscoring the need to focus on other factors such as comorbidities and host age for understanding the heterogeneity in disease outcomes for SARS-CoV-2.

## Introduction

The efficacy of masks in reducing infection risk has been shown in the context of SARS-CoV-2^1^ and for other respiratory viruses^2^. Masks reduce the amount of virus shed in exhaled breath of infected individuals^3^, reducing viral transmission potential. Although less effective than ‘source control’^1,2^, masking on the part of a contact also reduces infection risk. During the COVID-19 pandemic, an additional benefit of masking has been hypothesized: that masks can reduce disease severity in individuals who become infected despite masking^4–6^. The initial articles arguing that ‘masks do more’ than reduce viral transmission have been heavily cited^4,5,7^, with several follow-up articles that either express support of this hypothesis^6,8^ or are critical of it based on the extent of evidence presented^9–11^ or on findings from subsequent studies^12^.

The potential for masks to lower the risk of disease has been termed the ‘variolation hypothesis’^4^. The term variolation derives from a practice that was first documented in Asia in the 1500’s to reduce an individual’s risk of contracting smallpox. The practice involved inoculation of previously uninfected individuals using pulverized smallpox scabs. These challenged individuals only rarely developed severe disease in response to the inoculation but gained immunity to smallpox infection. A possible reason why variolation rarely led to severe disease is that the inoculum dose may have been low compared to a natural smallpox infection.

Here, we use mathematical modeling to develop a framework for assessing the plausibility of the variolation hypothesis as it pertains to respiratory viruses. We parameterize the models with estimated parameter values for SARS-CoV-2 and test the predictions of this framework using findings from existing experimental studies. Application of our framework to SARS-CoV-2 indicates that masking and other measures to reduce natural inoculum doses are highly unlikely to impact the risk of developing disease in those individuals who become infected. However, our findings indicate that these measures are effective at reducing infection risk, consistent with an existing body of empirical findings.

## Methods

### The relationship between inoculum dose and infection risk

The basic reproduction number R_0_ in epidemiology quantifies the expected number of secondary cases resulting from a single infected case in an otherwise susceptible host population^13^. Given a value of R_0_ and a measure of the extent of transmission heterogeneity between individuals, the probability of an emerging infectious agent establishing, rather than going stochastically extinct, can be calculated^14^. These epidemiological calculations have analogies at the within-host level, where the within-host basic reproduction number (*R*_0, within_) quantifies the expected number of cells that will become infected by virus progeny produced from a single infected cell early on in infection when target cells are readily available and host immune responses have not yet developed. We thus use mathematical expressions from the epidemiological literature to project how inoculum dose impacts the probability of an individual becoming infected (Supplemental Material). We parameterize the infection risk model based on existing SARS-CoV-2 literature estimates for *R*_0,_ within ^15^ and under a broad range of cell-to-cell heterogeneity levels. The range of heterogeneity levels we consider span from no cell-to-cell heterogeneity to extreme levels of heterogeneity with virus progeny from approximately 0.1% of infected cells being responsible for infecting 80% of the next generation of infected cells (Supplemental Material).

### The relationship between inoculum dose and the risk of developing severe disease

Within-host models are commonly used to understand viral load and immune response dynamics^16^. To project the relationship between inoculum dose and the risk of developing severe disease, we use an existing mathematical model for the viral and immune response dynamics during SARS-CoV-2 infection^15^. This model incorporates uninfected target cells, infected target cells, free virus, and the innate immune response. We extend this model to further incorporate an adaptive immune response, given the documented importance of the cellular immune response in clearing SARS-CoV-2 infection and in modulating disease severity^17^. We further add an equation to model tissue damage, which we ultimately use to quantify the extent of disease severity. Model equations and parameterizations are provided in the Supplemental Material. For any given parameterization (reflecting a given individual), we simulate the within-host model starting with different values for the initial number of infected cells.

### The inoculum dose in natural infections

To estimate the inoculum dose in natural SARS-CoV-2 infections, we used empirical estimates of the transmission bottleneck size of SARS-CoV-2^18–21^. The transmission bottleneck size *N*_b_ is defined as the number of viral particles that establish genetic lineages in an infected host. By contrast, inoculum dose is defined herein as the number of initially infected cells, which may be greater than *N*_b_. Under each considered inoculum dose, we analytically calculated the probability of *N*_b_ being 1, 2, 3, etc. viral particles, conditional on host infection (Supplemental Material). We used these probabilities to calculate the mean bottleneck size under any given inoculum dose. We compared these mean bottleneck sizes against the empirical mean estimate^18^ of N = 1.21 to determine a plausible range of natural inoculum doses.

## Results

### The expected relationship between inoculum dose and infection risk

To parameterize the infection risk model, we consider three values of *R*_0, within_: 7.4, 2.6, and 14.9, corresponding to the mean, low, and high estimates derived from a within-host viral dynamic model that was fit to viral load data from 17 infected individuals^15^. With *R*_0, within_ = 7.4 and in the absence of cell-to-cell heterogeneity (*k* = ∞), the risk of infection was close to 100% in the case of the inoculum dose being one or more initially infected cells (Figure 1). At any given inoculum dose, the risk of infection was lower at higher levels of cell-to-cell heterogeneity. However, even at extreme levels of cell-to-cell heterogeneity, infection risk saturated at 100% with inoculum doses of approximately 100-1000 initially infected cells. The overall patterns of infection risk were similar for a higher *R*_0, within_ (of 14.9) and a lower *R*_0, within_ (of 2.6). These results indicate that, for viral infections with high within-host basic reproduction numbers (*R*_0, within_>2), infection risk increases rapidly with increases in inoculum dose only over a range of low viral inoculum doses; at high inoculum doses, infection is already ensured.

**Figure 1.**
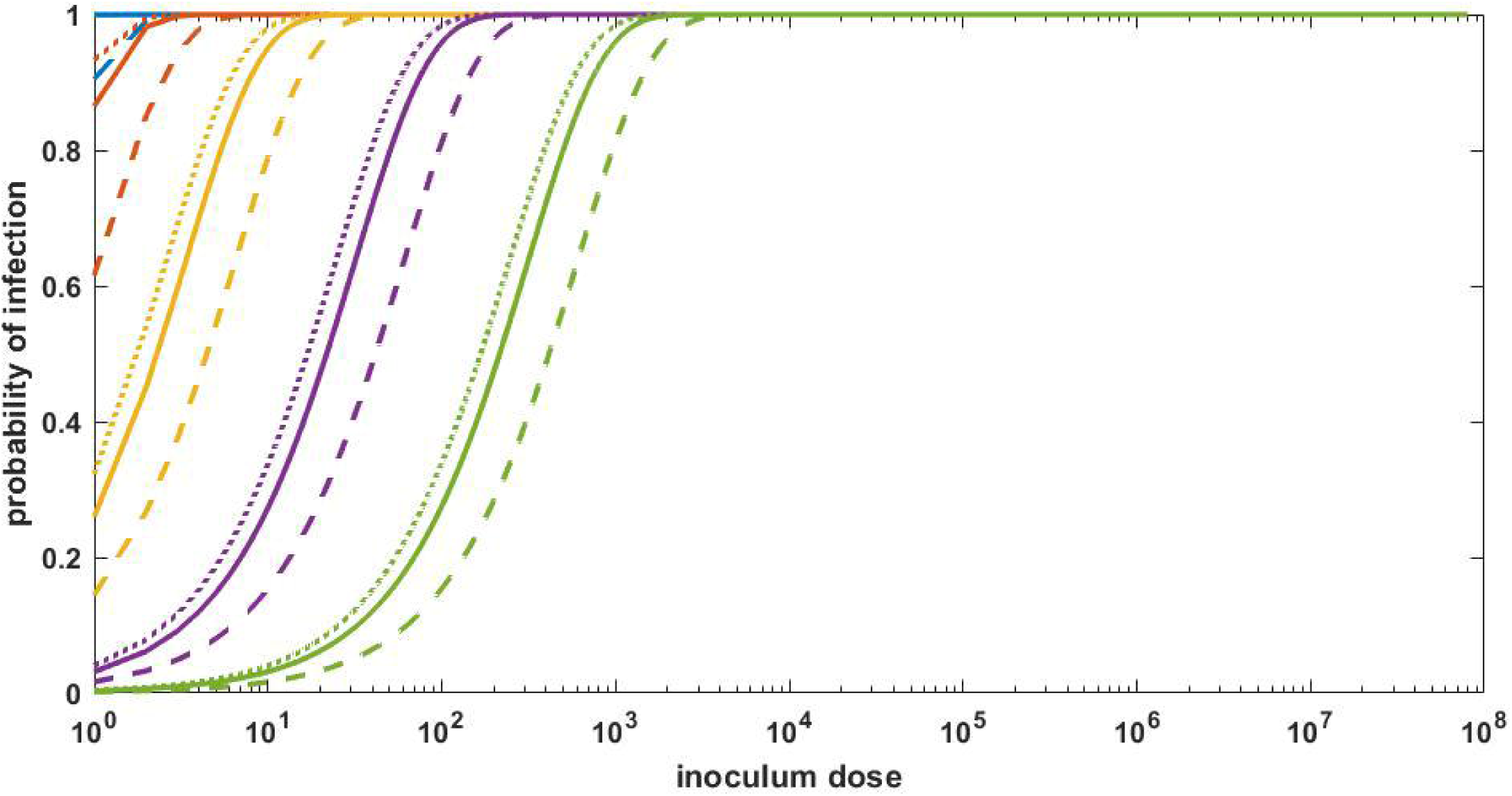
The relationship between inoculum dose and the risk of infection for SARS-CoV-2. Inoculum dose is defined here as the initial number of infected cells. Relationships are shown for *R*_0, within_ = 7.4 (solid lines), *R*_0, within_ = 2.6 (dashed lines), and *R*_0, within_ = 14.9 (dotted lines). Line colors denote the extent of cell-to-cell heterogeneity: overdispersion parameter *k* = ∞ (blue; no cell-to-cell heterogeneity), *k* = 1 (brown), *k* = 0.1 (yellow), *k* = 0.01 (purple), and *k* = 0.001 (green; extreme level of cell-to-cell heterogeneity). These *k* values correspond to approximately 65%, 40%, 10%, 1%, and 0.1% of the most infectious cells giving rise to 80% of secondary infected cells, respectively.

### The expected relationship between inoculum dose and risk of developing severe disease

Simulation of the within-host viral dynamic model, parameterized with baseline values and starting with an inoculum dose of 10 initially infected cells, recapitulated key features of SARS-CoV-2 within-host dynamics. Viral load increased over a period of approximately 6 days, peaked at approximately 10^7^ genome equivalents per ml, and then declined to undetectable levels within the following ∼8 days (**Figure 2A**). Tissue damage, driven by proinflammatory cytokines and T-cell-induced pathology, increased as viral load increased, peaked at around the same time as viral load, and then declined (**Figure 2B**). Only approximately 10% of target cells were killed over the course of the infection, with viral regulation resulting primarily from the innate immune response initially, followed by the cellular immune response (Supplemental Material).

**Figure 2.**
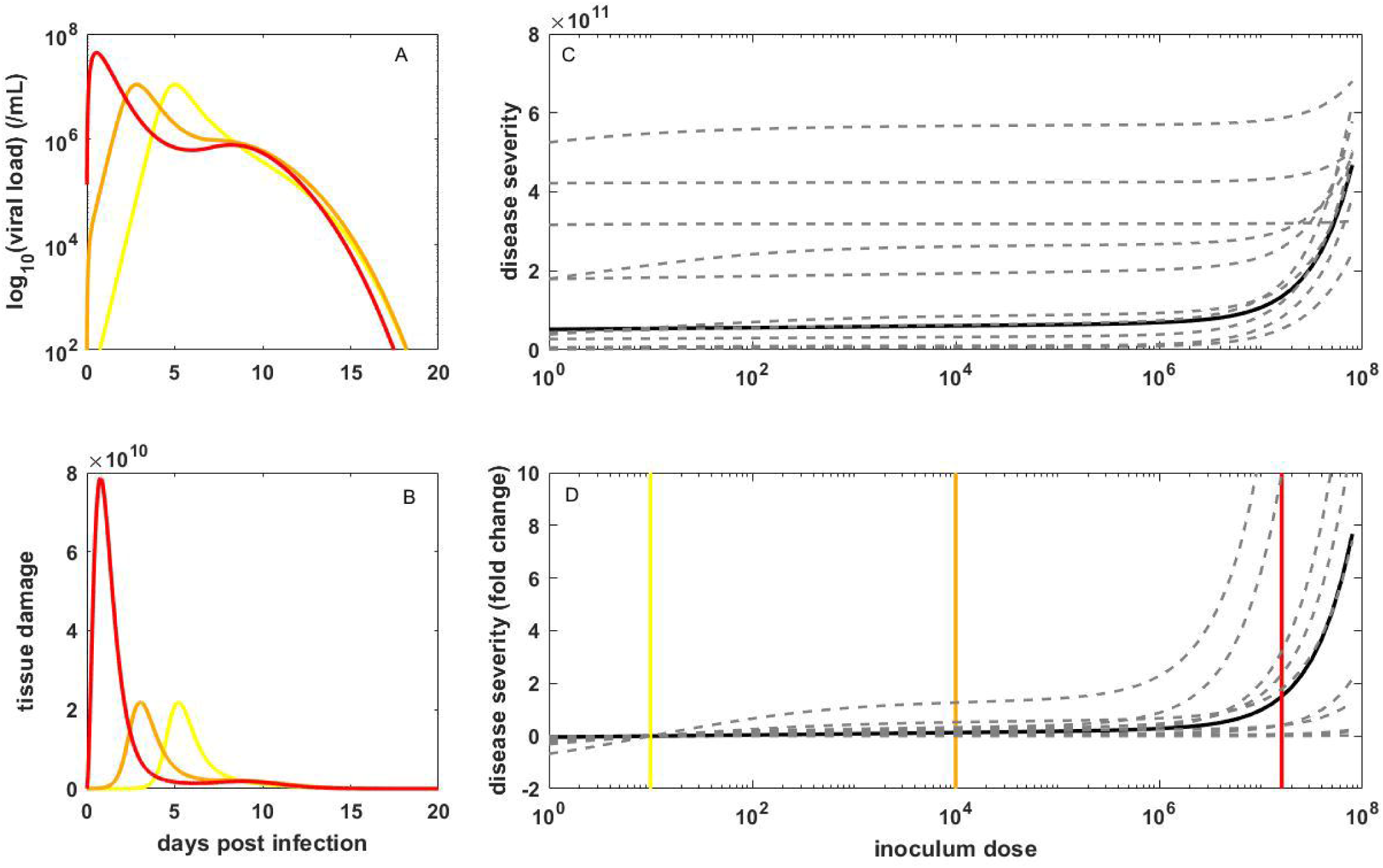
The relationship between inoculum dose and the risk of developing severe disease for SARS-CoV-2. (A) Viral dynamics, and (B) corresponding tissue damage dynamics, simulated using the within-host model parameterized with baseline values, for three inoculum doses: 10 (yellow), 10^4^ (orange), and 1.6×10^7^ (red) initially infected cells. (C) Disease severity of infection over a broad range of inoculum doses. Solid black line shows results for the within-host model parameterized at baseline values. Dashed gray lines show results for 10 individuals, with parameters sampled from the provided distributions detailed in the Supplemental Material. (D) Disease severity, for the baseline model parameterization (black) and 10 individuals (dashed gray), as in (C), in terms of fold change. Fold change is relative to the disease severity level resulting from infection starting with 10 infected cells. Vertical yellow, orange, and red lines in panels (C) and (D) show the inoculum doses used in the simulations shown in (A) and (B).

Increasing the inoculum dose by three orders of magnitude (10^4^ initially infected cells), decreased the time between infection and peak viral load (**Figure 2A**), and similarly sped up the dynamics of the other variables (**Figure 2B**; Supplemental Material). Despite these kinetic differences, the dynamics are quantitatively similar to those with a low inoculum dose of 10 initially infected cells such that the risk of developing severe disease was similar at these two doses. However, at an even higher inoculum dose of 1.6×10^7^ initially infected cells (corresponding to ∼20% of the total number of target cells), the within-host dynamics are substantially different from those at the two lower doses: viral titers peak at higher levels (**Figure 2A**), fewer target cells remain (Supplemental Material), and tissue damage is more substantial (**Figure 2B**), resulting in a higher risk of developing severe disease (**Figure 2C**).

To gauge the point at which the risk of developing severe disease increases, we simulated the within-host model across a wide range of inoculum doses, calculating the risk at each inoculum dose (**Figure 2C**). In **Figure 2D**, we plot this risk in terms of fold change relative to an infection starting from 10 initially infected cells. This figure shows that across >6 orders of magnitude difference in inoculum dose, from a single initially infected cell to ∼10^6^ initially infected cells, the risk of developing severe disease is insensitive to dose; only at extremely high doses does the risk of developing severe disease increase with an increase in dose.

To examine interindividual variation in the risk of developing severe disease, we simulated the within-host model 10 times, with parameter values drawn from distributions with mean values given by the baseline parameterized model (Supplemental Material). Simulated viral dynamics and tissue damage dynamics were variable between these 10 simulations (Supplemental Material), resulting in highly variable risks of developing severe disease across individuals at a given inoculum dose (**Figure 2C**). However, plotting the risk for each of these 10 individuals relative to the risk under the assumption of 10 initially infected cells again indicates that the risk of developing severe disease is insensitive to the inoculum dose until doses approach very high levels (**Figure 2D**).

Our finding that the risk of developing severe disease is insensitive to dose across a broad range of inoculum doses, ranging from a single initially infected cell to ∼10^6^ initially infected cells (∼1% of initially available target cells), can be generally understood in the context of how host immunity responds to viral infection. At any inoculum dose, host immunity develops in response to viral infection. When inoculum doses are not extremely large, this immune response can effectively regulate viral dynamics. At extremely large inoculum doses, however, the host immune response does not have the ability to quickly regulate within-host viral dynamics, and as such, the number of infected cells is significantly higher. A higher number of infected cells results in higher interferon levels, which act to control the viral infection but also lead to higher levels of interferon-induced pathology. Simplifications of the within-host model we use here demonstrate this point (Supplemental Material).

#### Inference of inoculum dose in natural infections

**Figure 3A** shows the expected transmission bottleneck size under a range of inoculum doses, under the same set of values of *R*_0, within_ and cell-to-cell heterogeneity levels considered in Figure 1. Even at an extreme level of cell-to-cell heterogeneity (*k* = 0.001), the inoculum dose that yields an expected transmission bottleneck size of 1.21 does not exceed 124 initially infected cells. Even if transmission bottleneck sizes were an order of magnitude higher (∼10 viral particles), the inoculum dose would not exceed ∼3000 initially infected cells. These results indicate that the inoculum dose in natural SARS-CoV-2 infections is very low. In this range of inoculum doses, reductions in dose would be expected to decrease the risk of infection but not have an effect on the risk of developing severe disease. In **Figure 3B**, we show the distribution of transmission bottleneck sizes that has been previously inferred from empirical studies^18^. **Figures 3C-G** show that expected distributions of transmission bottleneck sizes under different cell-to-cell heterogeneity levels, parameterized with inoculum doses that yield mean *N*_b_ estimates that are closest to the value of 1.21, quantitatively reproduce the inferred empirical distribution.

**Figure 3.**
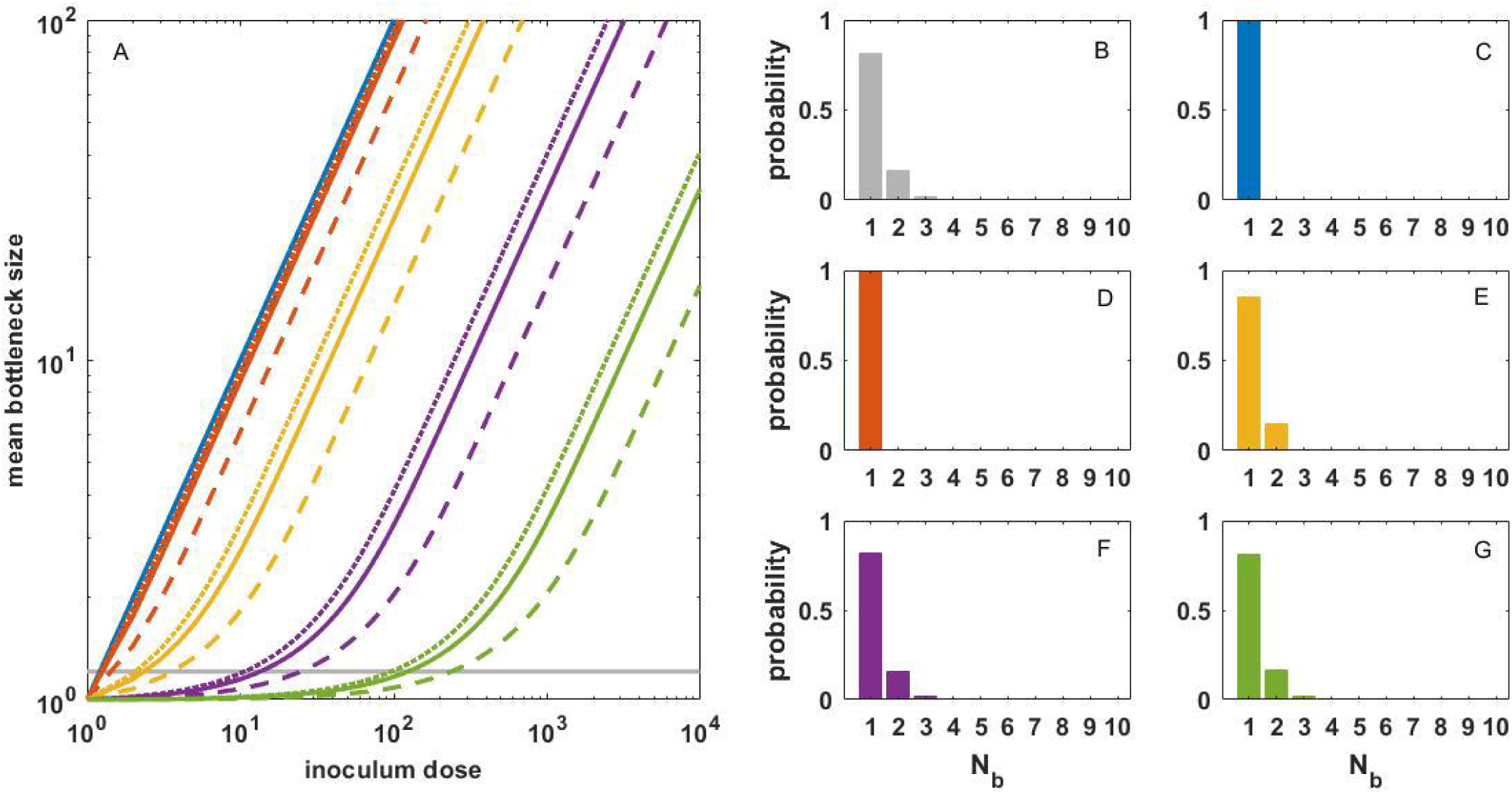
Inference of the natural inoculum dose. (A) The relationship between inoculum dose and mean bottleneck size. Lines show the expected relationship under a given within-host basic reproduction number and a given level of cell-to-cell heterogeneity. As in Figure 1, dashed, solid, and dotted lines correspond to *R*_0, within_ values of 2.6, 7.4, and 14.9, respectively, and line colors denote the extent of cell-to-cell heterogeneity. Gray line shows the mean transmission bottleneck size of 1.21. (B) Inferred distribution of transmission bottleneck sizes, reproduced from ^18^, showing the number of viral particles that establish genetic lineages in an infected individual and their corresponding probabilities. (C-G) Expected distribution of transmission bottleneck sizes for models parameterized with an *R*_0, within_ of 7.4 and inoculum doses that yield a mean bottleneck size that is closest to the previously inferred value of 1.21. Panels differ in their cell-to-cell heterogeneity levels. These doses correspond to 1 (C; *k* = Inf), 1 (D; *k* = 1), 2 (E; *k* = 0.1), 13 (F; *k* = 0.01), and 124 (G; *k* = 0.001) initally infected cells.

#### Analysis of experimental challenge studies

Our modeling provides two predictions relevant to the variolation hypothesis: (i) in the range of low inoculum doses, infection risk decreases with a decrease in inoculum dose, but the risk of developing severe disease (conditional on infection) is not substantially impacted; and (ii) in the range of very high inoculum doses, the risk of developing severe disease decreases with a decrease in inoculum dose, but infection risk is not impacted (individuals will become infected despite decreases in dose).

To test these predictions, we turn to experimental SARS-CoV-2 challenge studies. The most relevant of these studies are ones that measure disease outcomes in contact (sentinel) animals, across experimental designs that have the potential to modulate infection dose. In these experiments, we expect that inoculum doses of sentinel animals are of similar orders of magnitude to those of humans experiencing natural infection. We found two relevant studies^22,23^. The first study^22^ assessed the efficacy of masks for reducing transmission risk using a Syrian hamster model. They found that masking significantly reduced the risk of infection of sentinel hamsters from 10/15 (66.7%) to 6/24 (25.0%) (*p* = 0.018; Fisher exact test). The study also reported that the sentinel hamsters in the masked arms of the experiments had lower clinical severity scores and milder histopathological changes, consistent with the variolation hypothesis. However, the authors did not condition on infection and uninfected sentinel hamsters exhibited no clinical symptoms. Inclusion of uninfected sentinel hamsters therefore would bias clinical severity scores to be lower in the masked arms of the experiment relative to the unmasked arm of the experiment. We reanalyzed their data (Supplemental Material) and did not find a statistically significant difference in the clinical severity scores of infected sentinel hamsters between the masked and unmasked treatment groups (*p* = 0.07 for 5 dpi; *p* = 0.27 for 7 dpi; Mann-Whitney U test; Supplemental Material). The data from this study therefore indicate that masking reduces infection risk but do not demonstrate significant impact of masking on disease outcome during transmission from an index to a contact individual.

A second study^23^ examined SARS-CoV-2 transmission efficiency from inoculated to contact animals, also using the Syrian hamster model. The study found that transmission efficiency was high in exposures that lasted one or more hours when the index animals were inoculated with 1×10^4^ PFU of virus. In a follow-up experiment that examined transmission efficiency at different points in time following index inoculation, the authors allowed contact between sentinel and index cases for one to two hours, at different time periods post-inoculation. Transmission was found to be most efficient when viral load in the inoculated animal was high (17 h to 2 d post-inoculation), consistent with the risk of infection of a contact animal depending on dose when doses are low. However, infected contact animals across the different exposure time blocks did not exhibit statistically significant differences in infection severity as measured by weight loss (all p-values > 0.05; two-sample t-test; Supplemental Material), even though one would expect exposure during high viral load of the index case to increase inoculum dose. However, infected contact animals did exhibit a statistically significant (*p* = 0.004; two-sample t-test), yet small, amount of weight loss relative to their uninfected counterparts. These results are consistent with other experimental transmission studies on Syrian hamsters that found either small or insignificant amounts of weight loss in infected contact animals^24,25^.

Another set of studies that has the potential to give insight into the effect of dose on disease outcomes are those that modulate inoculum dose across a wide range of values in experimentally challenged donor animals. One such study^26^ assessed the effect of SARS-CoV-2 inoculum dose on seroconversion and fever development in a non-human primate model. Positive relationships were observed between deposited dose and seroconversion (an indicator of infection, albeit an imperfect one; **Figure 4A**) and also between deposited dose and fever development (**Figure 4B**). A similar effect was maintained when we reanalyzed the data by estimating the relationship between deposited dose and fever development, conditional on seroconversion (**Figure 4B**; Supplemental Material). As already remarked on in the original data analysis^26^, the median infectious dose that resulted in fever development was significantly higher than the median infectious dose that resulted in seroconversion (256 TCID50 vs. 52 TCID50), a result that was maintained when fever development was conditioned on seroconversion (**Figure 4**). We further fit an alternative model to these data to allow for a non-zero probability of developing fever at low inoculum doses, conditional on infection. The extended model, which was statistically preferred over the original logistic model, predicted an even higher inoculum dose for the median infectious dose that resulted in fever development (460 TCID50; **Figure 4B**). This analysis therefore provides empirical support for the modeling results presented above: at low inoculum doses, an increase in dose increases the risk of infection but not the risk of developing disease.

**Figure 4.**
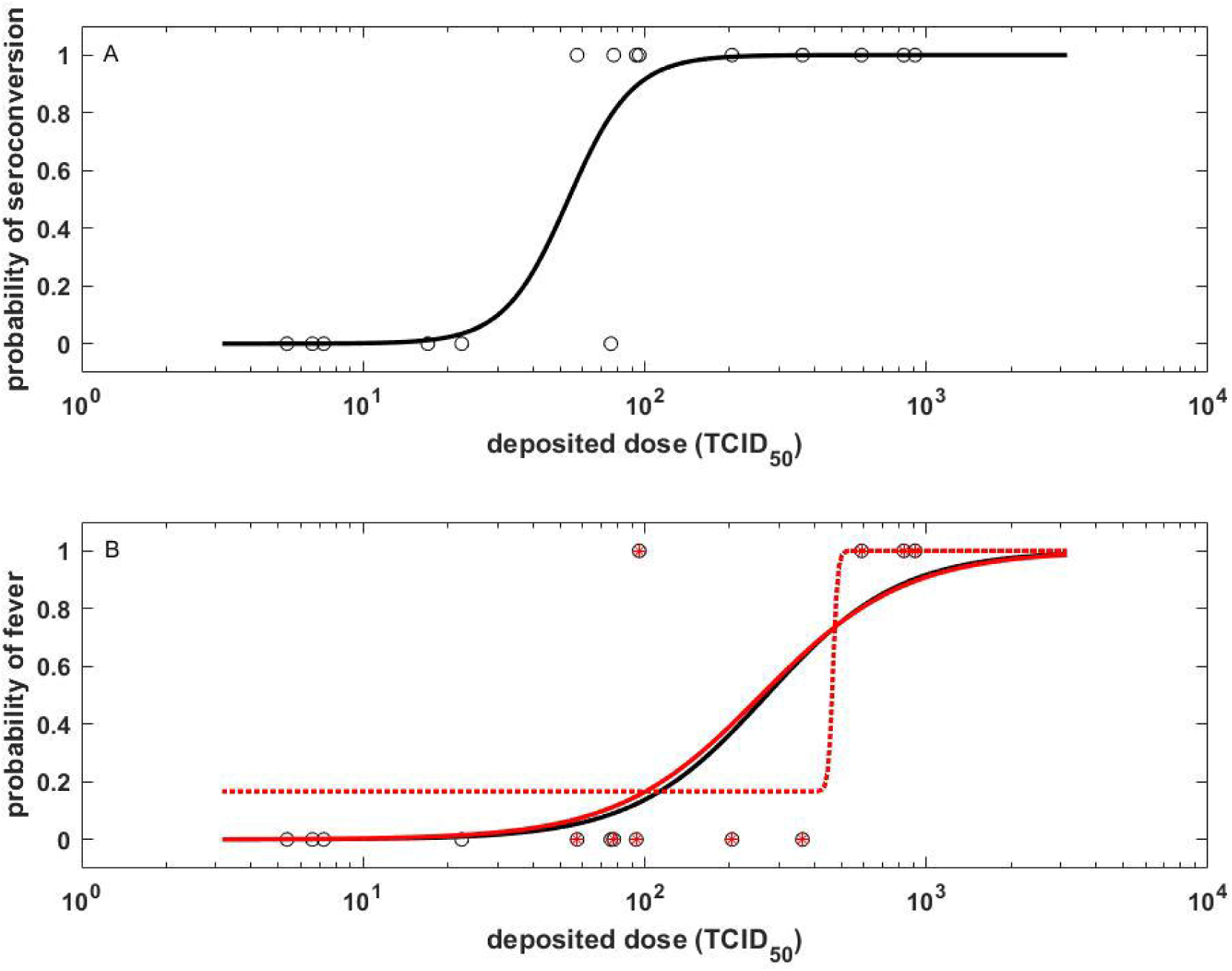
The relationship between inoculum dose, seroconversion, and fever development. Data derive from a non-human primate SARS-CoV-2 challenge study^26^, with inoculum doses ranging from low to high. Deposited doses are calculated from inoculum doses, incorporating deposition fraction estimates and accounting for variation in respiratory geometry. (A) The relationship between deposited dose and infection risk. Individual-level data points are shown as open black circles. Solid black line shows the fit of a logistic regression model. The median dose that results in seroconversion is 52 TCID50, as previously reported^26^. (B) The relationship between deposited dose and the probability of developing fever. Individual-level data points are shown as open black circles. Red asterisks within a subset of these data points indicate the subset of individuals that seroconverted. Solid black line shows the fit of a logistic regression model to all individual-level data points. Red solid line shows the fit of a logistic regression model to the subset of individuals that seroconverted. The median dose that results in fever development is is approximately 256 TCID50 for both models. The red dashed line shows the fit of the alternative logistic model to the subset of animals that seroconverted.

Another SARS-CoV-2 experimental challenge study examined disease outcomes at more than one dose^27^. However, this study considered dose ranges that ranged from high to very high (referred to as “low” and “high” dose, respectively, in their study) with all challenged animals becoming infected. At these dose levels, we expect that there might be a positive relationship between dose and disease severity. The findings from this study meet this expectation^27^ but do not provide support in favor of the variolation hypothesis because the inoculum doses used in this study lie outside the range of natural ones.

## Discussion

Here, we have used mathematical models to study the relationships between inoculum dose and the risks of infection and of developing severe disease. Based on parameterizations of these models for SARS-CoV-2, we argue that decreases in the inoculum dose, for example through masking, will only result in lower probabilities of infection when the natural inoculum dose is low. We further argue that decreases in the inoculum dose will only result in less severe disease when the natural inoculum dose is high. Our comparison of expected and empirical estimates of transmission bottleneck size indicates that natural inoculum doses are very low, such that masking (as documented^1^) is expected to reduce transmission potential. However, this means that masking is highly unlikely to reduce the risk of developing severe disease, conditional on infection. Our results thereby undermine the plausibility of the variolation hypothesis. Our results are consistent with experimental challenge studies, which have not found a significant difference in disease outcomes in contact animals infected with different but small inoculum doses; however, disease outcomes in index animals that are inoculated with high doses have been shown to differ, with higher doses resulting in more severe disease^27^.

We restricted our analyses to ones involving experimental challenge studies. While there are observational studies that have argued against the variolation hypothesis^12^, we feel that these studies offer an absence of evidence rather than evidence of absence, owing to uncertainty in the data. Other studies have been invoked to instead support the variolation hypothesis. For example, it has been argued that the higher asymptomatic rate on board of the Greg Mortimer ship destined for Antarctica (81%)^28^ relative to the asymptomatic rate on board of the Diamond Princess (17.9%)^29^ was due to masking on the Greg Mortimer^5^. However, alternative explanations, such as differences in the age distribution of the passengers or differences in the SARS-CoV-2 tests used, were not considered.

An alternative hypothesis to consider is that masks may modulate disease severity not by decreasing inoculum dose but by modulating the mode of transmission^30^. A viral inoculum delivered in larger droplets, such as those that comprise a spray, would become trapped in the nasal passages and upper airways. Conversely, small aerosolized particles can penetrate the lower lungs, where infection is more likely to result in severe symptoms. While masks that create a seal around the nose and mouth can limit inhalation of aerosols, most are more effective at limiting transfer of a droplet spray. Thus, while masking is expected to lower the overall number of infections, it could increase the proportion of cases resulting from inhalation directly to the lower respiratory tract. For this reason, we suggest that an effect of masking on modes of transmission is not consistent with the variolation hypothesis.

Our finding that masking and measures of social distancing that reduce inoculum dose are unlikely to do more than protect against infection has relevance to other respiratory viruses such as influenza, which is also characterized by a small transmission bottleneck size^31^. Masking would be expected to reduce incidence of infection, helping to limit the impact of influenza at a population level. To reduce disease in those infected, whether it be with influenza, SARS-CoV-2, or another respiratory virus characterized by a small inoculum dose, vaccination likely remains the most effective countermeasure^32,33^.

## Data Availability

All data produced in the present work are contained in the manuscript

## Acknowledgments

We thank Dr. Jasper Chan and Dr. Kwok-Yung Yuen for providing their study’s individual-level clinical severity score data upon request. We further thank Ketaki Ganti and Lucas Ferreri for providing individual-level weight data from the Ganti et. al. study, and Narendra Dixit for helpful conversations about their SARS-CoV-2 within-host model. This work was funded by NIH/NIAID Centers of Excellence in Influenza Research and Response (CEIRR), contract number 75N93021C00017 to KK, RA and ACL. DW was supported by the Simons Foundation via a Mathematical Modeling of Living Systems Investigator award, the Sloan Foundation via a Research Fellowship, the NSF via CAREER award PHY-2146260, the Gordon and Betty Moore Foundation grant 2919.02, and the Kavli Institute for Theoretical Physics by NSF grant PHY-1748958.

## References

1 Howard J, Huang A, Li Z, et al. An evidence review of face masks against COVID-19. Proc Natl Acad Sci USA 2021; 118: e2014564118.

2 Cowling BJ, Zhou Y, Ip DKM, Leung GM, Aiello AE. Face masks to prevent transmission of influenza virus: a systematic review. Epidemiol Infect 2010; 138: 449–56.

3 Leung NHL, Chu DKW, Shiu EYC, et al. Respiratory virus shedding in exhaled breath and efficacy of face masks. Nat Med 2020; 26: 676–80.

4 Gandhi M, Rutherford GW. Facial Masking for Covid-19 — Potential for “Variolation” as We Await a Vaccine. N Engl J Med 2020; 383: e101.

5 Gandhi M, Beyrer C, Goosby E. Masks Do More Than Protect Others During COVID-19: Reducing the Inoculum of SARS-CoV-2 to Protect the Wearer. J GEN INTERN MED 2020; published online July 31. DOI:10.1007/s11606-020-06067-8.

6 Sehrawat S, Rouse BT. COVID-19: disease, or no disease? - that is the question. It’s the dose stupid! Microbes and Infection 2021; 23: 104779.

7 Spinelli MA, Glidden DV, Gennatas ED, et al. Importance of non-pharmaceutical interventions in lowering the viral inoculum to reduce susceptibility to infection by SARS-CoV-2 and potentially disease severity. The Lancet Infectious Diseases 2021; 21: e296–301.

8 Van Damme W, Dahake R, van de Pas R, Vanham G, Assefa Y. COVID-19: Does the infectious inoculum dose-response relationship contribute to understanding heterogeneity in disease severity and transmission dynamics? Medical Hypotheses 2021; 146: 110431.

9 Trunfio M, Calcagno A, Bonora S, Di Perri G. Lowering SARS-CoV-2 viral load might affect transmission but not disease severity in secondary cases. The Lancet Infectious Diseases 2021; 21: 914–5.

10 Escandón K, Rasmussen AL, Bogoch II, et al. COVID-19 false dichotomies and a comprehensive review of the evidence regarding public health, COVID-19 symptomatology, SARS-CoV-2 transmission, mask wearing, and reinfection. BMC Infect Dis 2021; 21: 710.

11 Facial Masking for Covid-19. N Engl J Med 2020; 383: 2092–4.

12 Trunfio M, Longo BM, Alladio F, et al. On the SARS-CoV-2 “Variolation Hypothesis”: No Association Between Viral Load of Index Cases and COVID-19 Severity of Secondary Cases. Front Microbiol 2021; 12: 646679.

13 Anderson RM, May RM. Infectious diseases of humans: dynamics and control, Reprinted. Oxford: Oxford Univ. Press, 2010.

14 Lloyd-Smith JO, Schreiber SJ, Kopp PE, Getz WM. Superspreading and the effect of individual variation on disease emergence. Nature 2005; 438: 355–9.

15 Ke R, Zitzmann C, Ho DD, Ribeiro RM, Perelson AS. In vivo kinetics of SARS-CoV-2 infection and its relationship with a person’s infectiousness. Proc Natl Acad Sci USA 2021; 118: e2111477118.

16 Perelson AS. Modelling viral and immune system dynamics. Nat Rev Immunol 2002; 2: 28–36.

17 Moss P. The T cell immune response against SARS-CoV-2. Nat Immunol 2022; 23: 186–93.

18 Martin MA, Koelle K. Comment on “Genomic epidemiology of superspreading events in Austria reveals mutational dynamics and transmission properties of SARS-CoV-2”. Sci Transl Med 2021; 13: eabh1803.

19 Lythgoe KA, Hall M, Ferretti L, et al. SARS-CoV-2 within-host diversity and transmission. Science 2021; 372: eabg0821.

20 Braun KM, Moreno GK, Halfmann PJ, et al. Transmission of SARS-CoV-2 in domestic cats imposes a narrow bottleneck. PLoS Pathog 2021; 17: e1009373.

21 Braun K, Moreno G, Wagner C, et al. Limited within-host diversity and tight transmission bottlenecks limit SARS-CoV-2 evolution in acutely infected individuals. Evolutionary Biology, 2021 DOI:10.1101/2021.04.30.440988.

22 Chan JF-W, Yuan S, Zhang AJ, et al. Surgical Mask Partition Reduces the Risk of Noncontact Transmission in a Golden Syrian Hamster Model for Coronavirus Disease 2019 (COVID-19). Clinical Infectious Diseases 2020; 71: 2139–49.

23 Ganti K, Ferreri LM, Lee C-Y, et al. Timing of exposure is critical in a highly sensitive model of SARS-CoV-2 transmission. PLoS Pathog 2022; 18: e1010181.

24 Port JR, Yinda CK, Avanzato VA, et al. Increased small particle aerosol transmission of B.1.1.7 compared with SARS-CoV-2 lineage A in vivo. Nat Microbiol 2022; 7: 213–23.

25 Port JR, Yinda CK, Owusu IO, et al. SARS-CoV-2 disease severity and transmission efficiency is increased for airborne compared to fomite exposure in Syrian hamsters. Nat Commun 2021; 12: 4985.

26 Dabisch PA, Biryukov J, Beck K, et al. Seroconversion and fever are dose-dependent in a nonhuman primate model of inhalational COVID-19. PLoS Pathog 2021; 17: e1009865.

27 Imai M, Iwatsuki-Horimoto K, Hatta M, et al. Syrian hamsters as a small animal model for SARS-CoV-2 infection and countermeasure development. Proc Natl Acad Sci USA 2020; : 202009799.

28 Ing AJ, Cocks C, Green JP. COVID-19: in the footsteps of Ernest Shackleton. Thorax 2020; 75: 693–4.

29 Mizumoto K, Kagaya K, Zarebski A, Chowell G. Estimating the asymptomatic proportion of coronavirus disease 2019 (COVID-19) cases on board the Diamond Princess cruise ship, Yokohama, Japan, 2020. Eurosurveillance 2020; 25. DOI:10.2807/1560-7917.ES.2020.25.10.2000180.

30 Prather KA, Wang CC, Schooley RT. Reducing transmission of SARS-CoV-2. Science 2020; 368: 1422–4.

31 McCrone JT, Woods RJ, Martin ET, Malosh RE, Monto AS, Lauring AS. Stochastic processes constrain the within and between host evolution of influenza virus. eLife 2018; 7. DOI:10.7554/eLife.35962.

32 Gross PA. The Efficacy of Influenza Vaccine in Elderly Persons: A Meta-Analysis and Review of the Literature. Ann Intern Med 1995; 123: 518.

33 Feikin DR, Higdon MM, Abu-Raddad LJ, et al. Duration of effectiveness of vaccines against SARS-CoV-2 infection and COVID-19 disease: results of a systematic review and meta-regression. The Lancet 2022; 399: 924–44.

