## Supplemental Material for "Masks Do No More Than Prevent Transmission: Theory and Data Undermine the Variolation Hypothesis"

Supplemental Material

Sections

I. *Calculations for projecting infection risk*
II. *Within-host model equations and parameterizations*
III. *Within-host model simplifications that demonstrate the reason for insensitivity of disease severity to inoculum dose at low doses*
IV. *Calculation of transmission bottleneck sizes*
V. *Reanalysis of Chan et al. (2020) Clinical Infectious Diseases*
VI. *Analysis of weight loss data in contact animals from Ganti et al. (2022) PLoS Pathogens*
VII. *Further analysis of Dabisch et al. (2021) PLoS Pathogens*
I. *Calculations for projecting infection risk*

We use a branching process approach to calculate the probability of an individual becoming infected, given an initial number of infected cells. This initial number corresponds to the number of cells infected via inoculum virus, rather than through local within-host rounds of replication. To calculate infection risk using this approach, we need to specify an “offspring distribution”: a distribution that quantifies the number of infected cells a single infected cell generates. We further need to parameterize this distribution. Based on previous studies using branching processes to quantify probabilities of disease emergence following spillover^1^, we use a negative binomial distribution, parameterized with a mean and overdispersion parameter *k*. In the context of within-host infection establishment, the mean of this distribution corresponds to the within-host basic reproduction number *R*_0, within_. The overdispersion parameter ranges from 0 to ∞, with lower values of *k* corresponding to more cell-to-cell heterogeneity.

To ensure the robustness of our results, we consider multiple different parameterizations of this negative binomial distribution. We consider three different values for *R*_0, within_: 7.4, 2.6, and 14.9. *R*_0, within_ = 7.4 corresponds to the mean value of *R*_0, within_ estimated for SARS-CoV-2^2^. *R*_0, within_ = 2.6 and *R*_0, within_ = 14.9 correspond to the lowest and the highest values of *R*_0, within_ estimated in this study. We also consider five different values for the overdispersion parameter *k*: *k* = ∞ (corresponding to no cell-to-cell heterogeneity and resulting in a Poisson distribution for offspring), *k* = 1 (resulting in a geometric distribution for offspring), *k* = 0.1, *k* = 0.01, and *k* = 0.01 (corresponding to extreme cell-to-cell heterogeneity in offspring numbers). With three different *R*_0, within_ values and five different *k* values, we thus consider 15 different negative binomial parameterizations that span the range of biologically plausible offspring distributions (Figure S1).

The probability of an infection establishing within a host (rather than going stochastically extinct) is given by:

$$p_{\mathrm{infection}}=1-p_{\mathrm{extinction},1}^{n}$$

where $p_{\mathrm{extinction},1}$is the probability that the lineage stemming from an initially infected cell goes stochastically extinct, and the exponent *n* is the inoculum dose (operationally defined as the initial number of infected cells). The value of $p_{\mathrm{extinction},1}$ is calculated numerically using a branching process approach, under a parameterized offspring distribution. For a negative binomial offspring distribution, the probability generating function is given by^1^:

$$g\left( s \right)=\left[ 1+\frac{R_{0,\mathrm{within}}}{k}(1-s) \right]^{-k}$$

The solution to $p_{\mathrm{extinction},1}$ is given by the unique solution $q=g\left( q \right)$ on the interval (0,1). We confirmed our branching process results through stochastic simulations.

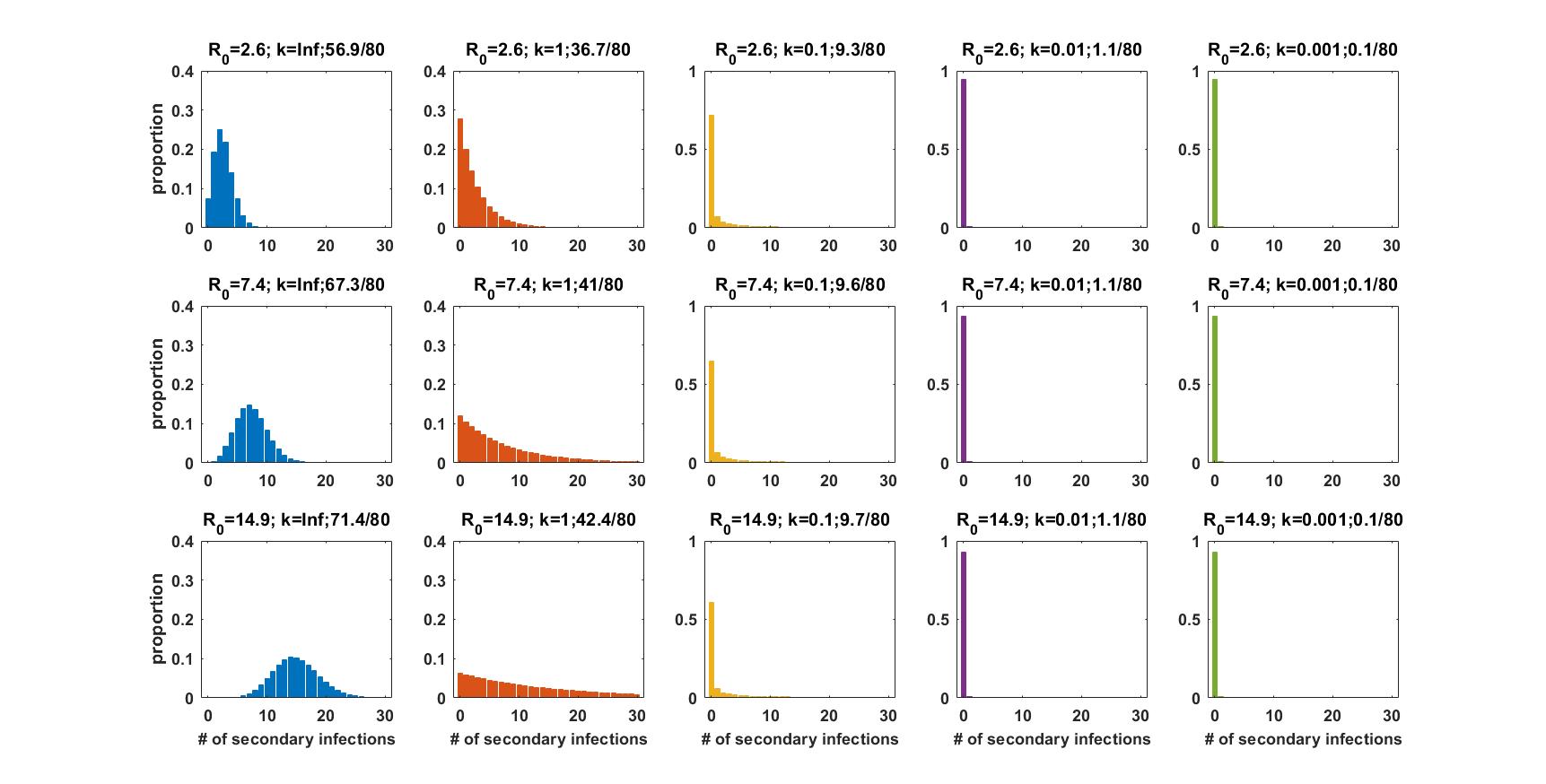

**Figure S1. Offspring distributions considered in the calculation of infection risk.** Rows correspond to the low, mean, and high *R*_0, within_ values considered. Columns correspond to the level of cell-to-cell heterogeneity considered, ranging from no cell-to-cell heterogeneity (*k* = ∞) to extreme heterogeneity (*k* = 0.001), according to the same color scheme as Figure 1 in the main text. Each panel title includes the distribution’s parameterization, consisting of *R*_0, within_ and the overdispersion parameter *k*. Each panel title further includes the percentage of cells that are responsible for generating 80% of secondary infected cells (written as *x*/80, where the percentage is *x*). This percentage is commonly used in the epidemiological literature to report the extent of transmission heterogeneity. We calculated this percentage using a previously described approach^3^.

II. *Within-host model equations and parameterizations*

The within-host model formulation we use builds on a recently published model that was parameterized by a combination of empirical estimates and fits to viral load data from SARS-CoV-2 infected individuals^2^. Based on existing within-host model structures for acute respiratory infections such as influenza^4^, the published SARS-CoV-2 model is given by^2^:

$${dT}/{dt=-\beta VT-\Phi IT+\rho R}$$

$${dR}/{dt=\Phi IT-\rho R}$$

$${dE}/{dt=\beta VT-kE}$$

$${dI}/{dt=kE-\delta I}$$

$${dV}/{dt=\pi I-cV}$$

The variables correspond to:

*T* = uninfected target cells that are susceptible to viral infection;

*R* = uninfected target cells that are refractory to viral infection due to being in an anti-viral state;

*E* = infected cells that are not yet producing viral progeny (that is, cells in their eclipse phase);

*I* = infected cells that are actively producing viral progeny;

*V* = free virus.

The number of uninfected target cells that are susceptible to viral infection *T* decreases with cells becoming infected and with cells entering a refractory anti-viral state, and increases with refractory cells becoming re-susceptible to infection. The number of refractory cells *R* increases with cells entering a refractory anti-viral state, and decreases with refractory cells becoming re-susceptible to infection. The number of eclipse phase cells *E* increases with cells becoming infected and decreases with cells transitioning to being productively infected. The number of actively producing infected cells *I* increases with cells transitioning to being productively infected and decreases with infected cell death. The concentration of free virus *V* increases with viral progeny produced from productively infected cells and decreases with viral clearance. In this model, innate immune mediators, such as interferons, are not explicitly modeled, but are assumed to be in quasi-steady-state with productively infected cells, such that their concentration at any point in time is proportional to *I*. The term $\Phi IT$ thus represents susceptible target cells becoming refractory to infection by contact with innate immune mediators (rather than productively infected cells per se).

We further phenomenologically incorporate the adaptive (cellular) immune response, based on a previously described approach^4^, by letting the infected cell death rate *δ* depend on the time since infection. Specifically, we let $\delta(t)=\delta_{I}$ when $t<\mu$, and $\delta(t)=\delta_{I}e^{\sigma(t-\mu)}$ when $t\geq\mu$, where *t* denotes the time since infection and the parameter *μ* quantifies the time at which the T-cell immune response starts. We further extend the model to simulate tissue damage:

$${dD}/{dt=\alpha I\Lambda+\omega I-\gamma D}$$

This tissue damage equation is based on existing equations used for simulating symptom development and tissue damage in within-host models^5–7^. The variable $\Lambda$ represents relative T-cells levels, with $\Lambda(t)=1$ when $t<\mu$, and $\Lambda=e^{\sigma(t-\mu)}$ when $t\geq\mu$. The equation captures tissue damage arising from the killing of infected cells by T-cells and from proinflammatory cytokines, and resolving at a constant rate. Finally, as in ^7^, we summarize the severity of an infection by integrating over tissue damage over the course of an infection:

$$disease severity= \int D\left( t \right)dt$$

Our baseline parameterization of this within-host model is provided in Table S1. Figure S2 shows the dynamics of the model variables not shown in Figure 2 (*T*, *R*, *E*, *I*).

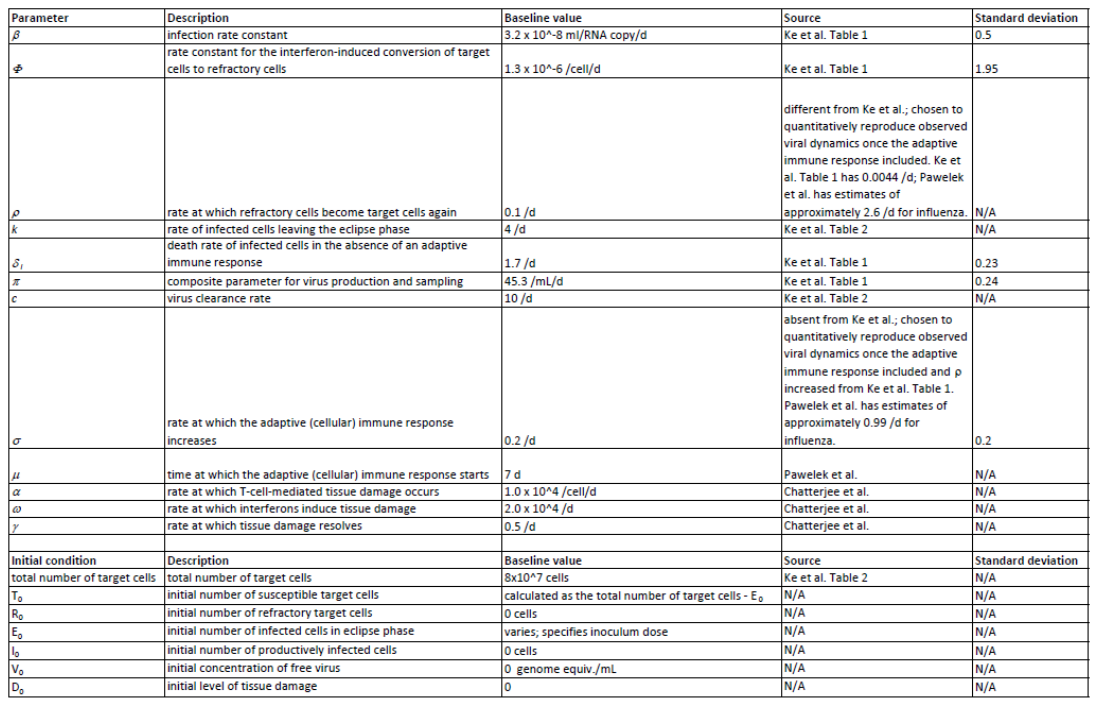

**Table S1.** Model parameters and initial conditions. Baseline model parameter values are provided, along with their sources. Specified sources include Ke et al.^2^, Pawelek et al.^4^, and Chatterjee et al.^7^. For clarity, parameter descriptions are taken directly from source papers, where possible. Parameter standard deviations used in the modeling of interindividual variation are also provided. As in Ke et al.^2^, these standard deviations are standard deviations of lognormal distributions.

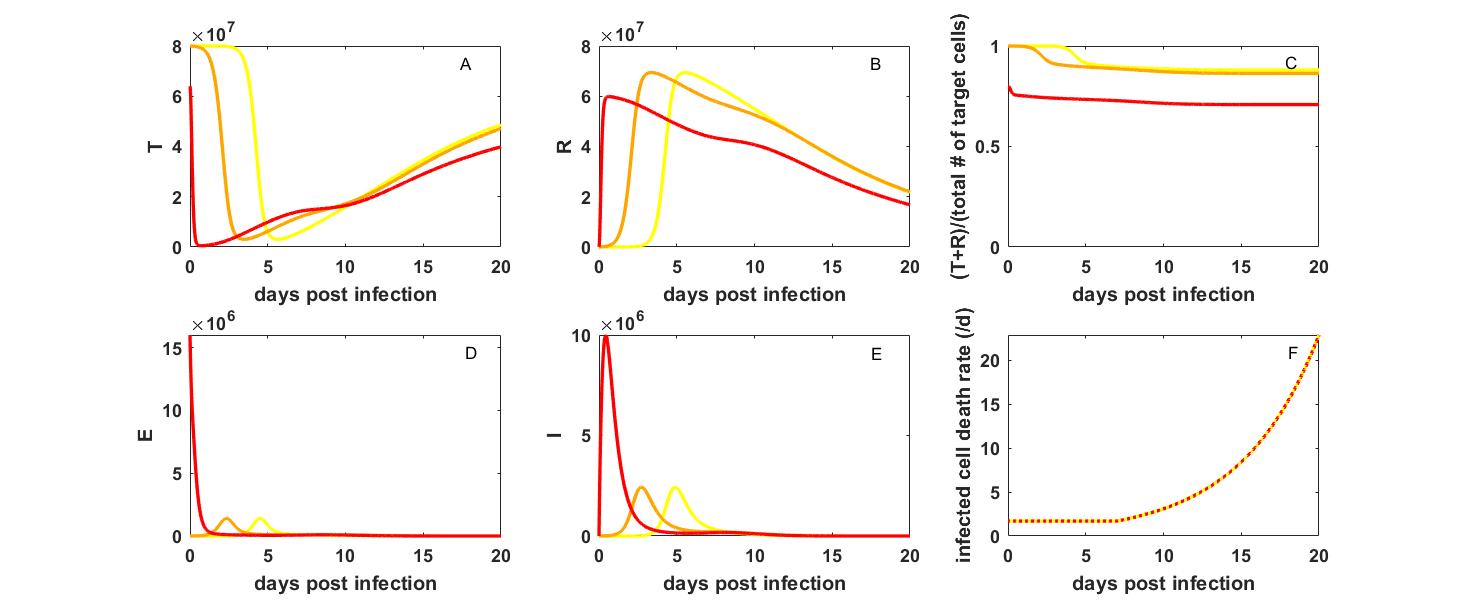

**Figure S2. Dynamics of the within-host model variables not shown in Figure 2.** (A) The dynamics of uninfected cells that are susceptible to infection. (B) The dynamics of uninfected cells that are refractory to infection. (C) The fraction of total target cells that are uninfected. (D) The number of eclipse-phase infected cells. (E) The number of productively infected cells. (F) The infected cell death rate *δ*. Analogous to the color-coding in Figure 2 of the main text, yellow, orange, and red lines correspond to inoculum doses of 10, 10^^4^, and 1.6x10^7^ initially infected cells, respectively.

Variation around the baseline model parameterization is introduced by sampling a subset of the model parameter values from log-normal distributions with the means given by the baseline parameterization and specified standard deviations (Table S1). Specifically, we consider interindividual variation in the parameters *β*, *δ*_I_, *π*, *Φ*, and *σ*. Variation in *Φ* reflects interindividual variation in the strength of the innate immune response. Variation in *σ* reflects interindividual variation in the speed at which the cellular immune response occurs. Variation in the other parameters reflects interindividual variation arising from other host factors. Figure S3 shows the viral load dynamics and the tissue damage dynamics from 10 individuals, with the values for each of these 5 parameters drawn randomly from their specified log-normal distributions.

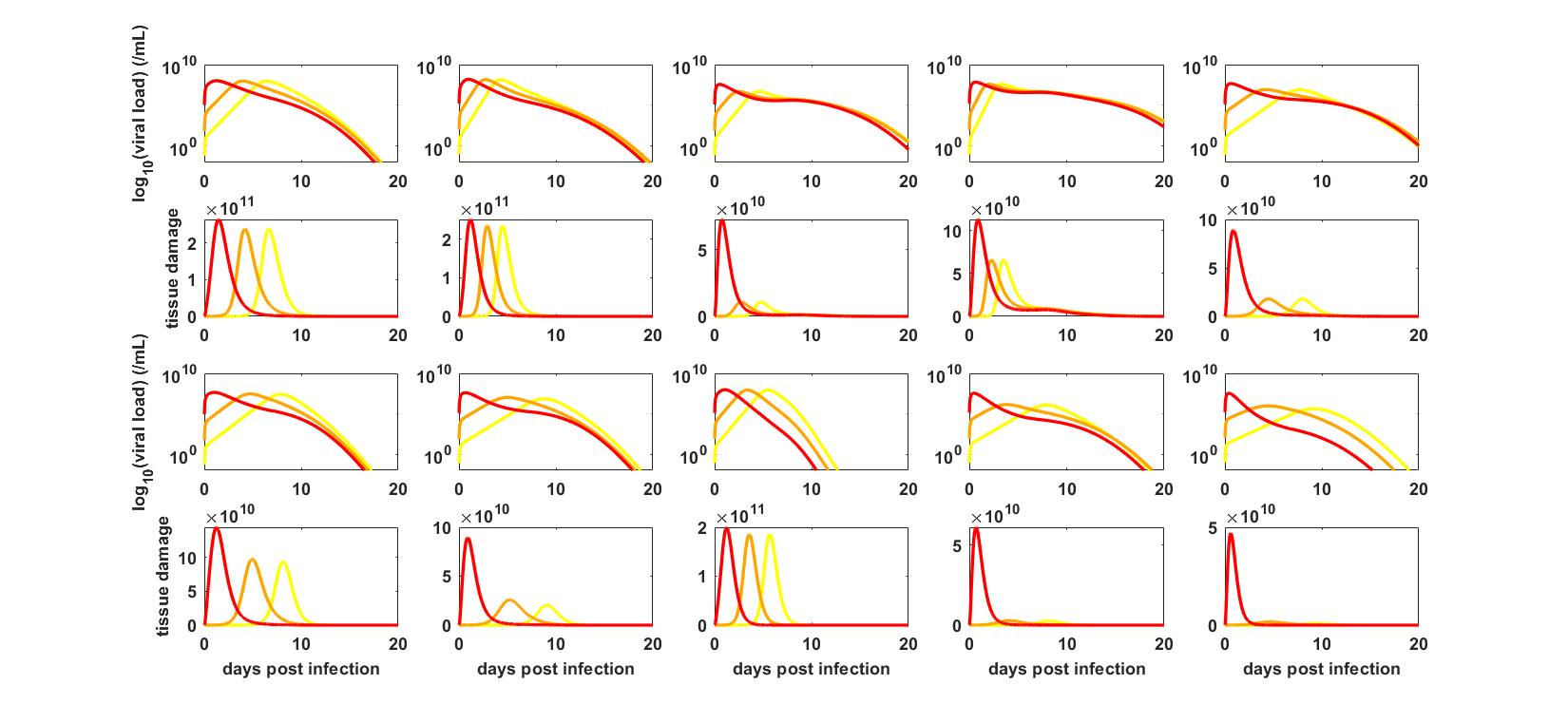

**Figure S3. Viral load and tissue damage dynamics of 10 individuals.** (First row) Viral load dynamics of individuals 1-5. (Second row) Corresponding tissue damage dynamics of individuals 1-5. (Third row) Viral load dynamics of individuals 6-10. (Fourth row) Corresponding tissue dynamics of individuals 6-10. Colored lines in each panel correspond to inoculum doses of 10 (yellow), 10^4^ (orange), and 1.6x10^7^ (red) initially infected cells.

III. *Within-host model simplifications that demonstrate the reason for insensitivity of disease severity to inoculum dose at low to moderate doses*

As described in the above section, we model tissue damage according to the following equation:

$${dD}/{dt=\alpha I\Lambda+\omega I-\gamma D}$$

This equation is based on formulations in previously published studies, and includes the roles that T-cells and the innate immune response play in contributing to tissue damage and ultimately disease severity. To better understand why disease severity is insensitive to inoculum dose across a broad range of doses (from 1 initially infected cell to ~10^6^ initially infected cells), we can consider two separate simplifications of this equation. The first simplification is one in which we assume that only the innate immune response contributes to tissue damage: ${dD}/{dt=\omega I-\gamma D}$. In this case, patterns of disease severity across inoculum doses qualitatively recapitulate the patterns shown in Figure 2 of the main text: disease severity is largely insensitive to inoculum dose until very high dose levels are reached (**Figure S4**). Because T-cells do not contribute to tissue damage, absolute magnitudes of tissue damage are lower in Figure S4 than in Figure 2.

The second simplification is one in which we assume that only the T-cell response contributes to tissue damage: ${dD}/{dt=\alpha I\Lambda-\gamma D}$. In this case, patterns of disease severity across inoculum doses similarly qualitatively recapitulate the patterns shown in Figure 2 of the main text: disease severity is largely insensitive to inoculum dose until very high dose levels are reached (**Figure S5**). Because the innate immune response does not contribute to tissue damage, absolute magnitudes of tissue damage are lower in Figure S5 than in Figure 2. We can understand this pattern because in this case, up to day 7, T-cells are only present at low numbers ($\Lambda=1$), such that increases in tissue damage up to this time are proportional to the number of infected cells ($\alpha I$). At inoculum doses of <10^6^ initially infected cells, the dynamics of infected cells are quantitatively similar, with the kinetics just offset (see Figure S2E). As such, tissue damage dynamics will also be quantitatively similar, with kinetics just offset (Figure S5B). If we instead assume that background levels of T-cells do not contribute to tissue damage, we can rewrite the equation for tissue damage as ${dD}/{dt=\alpha I(\Lambda-1)-\gamma D}$. In this case, disease severity does not have a consistent or strong relationship with inoculum dose (**Figure S6**). The most prominent pattern in this case is a negative relationship between inoculum dose and disease severity. This can be understood in the context of viral kinetics: at higher inoculum doses, the viral infection is over earlier. As such, the number of infected cells after day 7 (once T-cells expand beyond their background levels) is lower at higher inoculum doses than at lower inoculum doses. Tissue damage, and associated disease severity, may therefore be lower at higher inoculum doses.

Together, **Figures S4-S6** indicate that our results shown in Figure 2C and 2D, which show that disease severity is insensitive to inoculum dose across a broad range of inoculum doses (1-approx. 10^6^ initially infected cells), are due to the ability of the host immune response to effectively regulate viral dynamics at these inoculum doses. At very high inoculum doses, however, the host immune response loses its ability to quickly regulate within-host viral dynamics, and as such, the number of infected cells is significantly higher, which results in higher interferon levels, and ultimately a higher level of interferon-induced pathology.

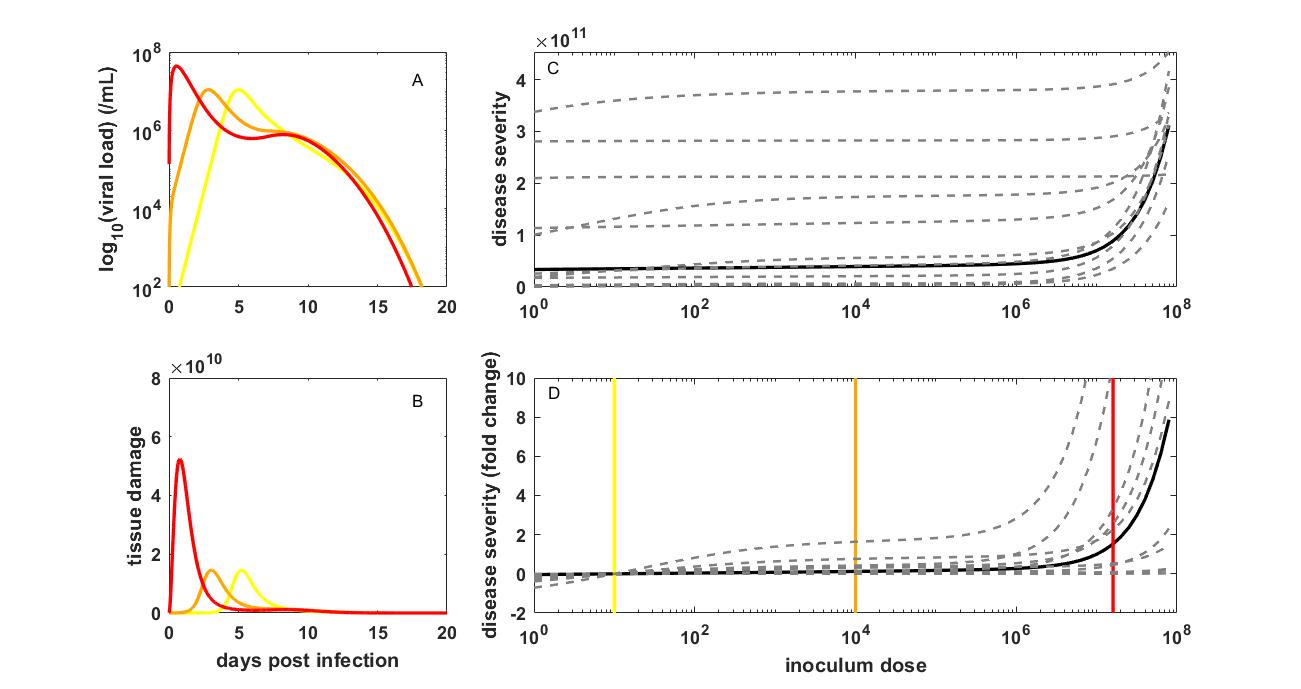

**Figure S4. The relationship between inoculum dose and the risk of developing severe disease for SARS-CoV-2 when only the innate immune response contributes to tissue damage.** (A) Viral dynamics, and (B) corresponding tissue damage dynamics, as in Figure 2, with the equation for tissue damage given instead by ${dD}/{dt=\omega I-\gamma D}$. (C) Disease severity of infection over a broad range of inoculum doses, as in Figure 2, with the equation for tissue damage instead given by ${dD}/{dt=\omega I-\gamma D}$. (D) Disease severity, for the baseline model parameterization (black) and 10 individuals (dashed gray), as in (C), in terms of fold change. Disease severity values used to calculated fold change are from panel (C).

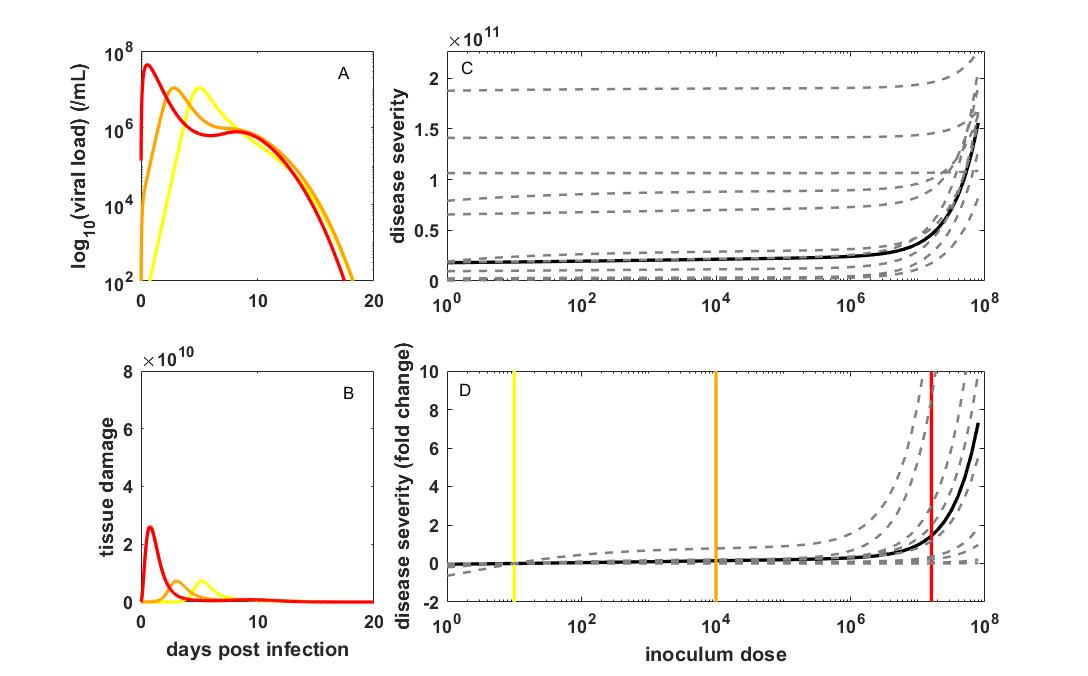

**Figure S5. The relationship between inoculum dose and the risk of developing severe disease for SARS-CoV-2 when only the adaptive (T-cell) response contributes to tissue damage.** (A) Viral dynamics, and (B) corresponding tissue damage dynamics, as in Figure 2, with the equation for tissue damage given instead by ${dD}/{dt=\alpha I\Lambda-\gamma D}$. (C) Disease severity of infection over a broad range of inoculum doses, as in Figure 2, with the equation for tissue damage instead given by ${dD}/{dt=\alpha I\Lambda-\gamma D}$. (D) Disease severity, for the baseline model parameterization (black) and 10 individuals (dashed gray), as in (C), in terms of fold change. Disease severity values used to calculated fold change are from panel (C).

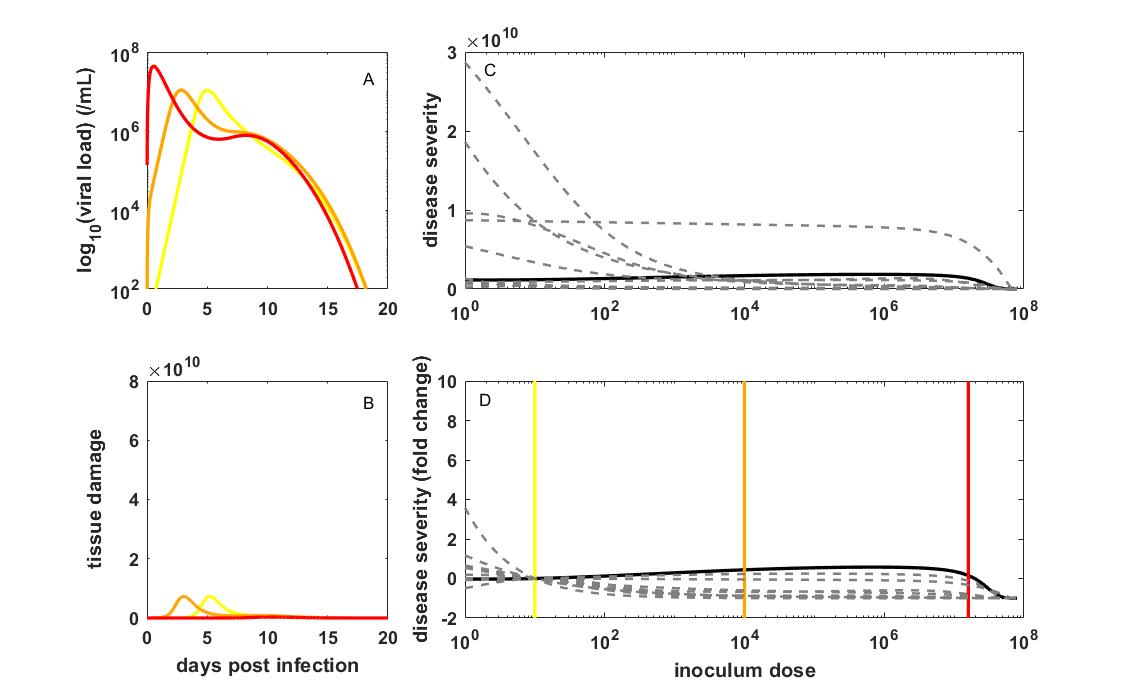

**Figure S6. The relationship between inoculum dose and the risk of developing severe disease for SARS-CoV-2 when only the adaptive (T-cell) response above background levels contributes to tissue damage.** (A) Viral dynamics, and (B) corresponding tissue damage dynamics, as in Figure 2, with the equation for tissue damage given instead by ${dD}/{dt=\alpha I(\Lambda-1)-\gamma D}$. (C) Disease severity of infection over a broad range of inoculum doses, as in Figure 2, with the equation for tissue damage instead given by ${dD}/{dt=\alpha I(\Lambda-1)-\gamma D}$. (D) Disease severity, for the baseline model parameterization (black) and 10 individuals (dashed gray), as in (C), in terms of fold change. Disease severity values used to calculated fold change are from panel (C).

IV. *Calculation of transmission bottleneck sizes*

For a given value of *R*_0, within_, a given level of cell-to-cell heterogeneity *k*, and a given inoculum dose, we can calculate the expected transmission bottleneck size as follows. We first calculate the probability that an infection establishes under a single initially infected cell, given *R*_0, within_ and *k*. This is given $p_{\mathrm{establishment},1}={1-p}_{\mathrm{extinction},1}$. The calculation of $p_{\mathrm{extinction},1}$ is detailed in Supplemental Material section I. For a given inoculum size of *N* initially infected cells, we can then calculate the probability that exactly *n* of these *N* infected cells establish a genetic lineage in the host. This is given by a binomial distribution, with *N* being the number of trials and $p_{\mathrm{establishment},1}$ being the probability of success, evaluated at *n*. Because transmission bottleneck sizes are calculated only in the case of infection, we need to condition on infection, such that the probability of the transmission bottleneck size being *n* (with *n*$\geq$*1*) is given by:

$${P(X=n;N, p}_{\mathrm{establishment},1})={Bin(n;N{,p}_{\mathrm{extinction},1})/(1-Bin\left( 0;N{,p}_{\mathrm{extinction},1} \right))}$$

The mean transmission bottleneck size is then given by:

$$N_{b}=\sum_{n=1}^{\infty} {nP(X=n;N, p}_{\mathrm{establishment},1})$$

We compare this mean transmission bottleneck size to inferred mean transmission bottleneck sizes. Existing studies have found that transmission bottleneck sizes are very small^8–11^, with the most quantitative estimate in humans indicating that between 1 and 3 viral particles establish genetic lineages within >99% of infected hosts^8^, with a mean *N*_b_ of 1.21.

V. *Reanalysis of Chan et al. (2020) Clinical Infectious Diseases*

We reanalyzed the clinical severity scores of the contact animals to determine whether masking in this experiment resulted in lower levels of clinical severity. We obtained the individual-level data upon request from the authors. We first asked whether clinical severity scores were significantly different between the unmasked (Experiment 1) contacts and the contacts in Experiment 2, 3, or both using a Mann-Whitney U test. The authors previously used a Student t-test on this dataset, but because of the integer-values severity scores and their deviation from a normal distribution, we instead used the more appropriate Mann-Whitney U test. We found, considering all contacts, regardless of whether they became infected or not, that clinical severity scores were significantly lower in the experiments that included masks than in the unmasked experiment, both at 5 dpi (p = 0.0167) and at 7 dpi (p = 0.0311) (Table S2). However, we then excluded the contacts that did not become infected from the analysis to determine whether clinical severity scores were different between the unmasked and masked experiments, conditional on infection. Table S3 shows that clinical severity scores for the masked hamsters that got infected were not significantly different from the unmasked hamsters that got infected, at either 5 dpi nor 7 dpi.

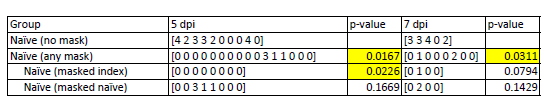

**Table S2.** Clinical scores of hamsters in the three experiments performed. Both uninfected and infected hamsters are included in the shown data. P-values show comparisons against the no mask experiment on the corresponding day post inoculation of the index hamster.

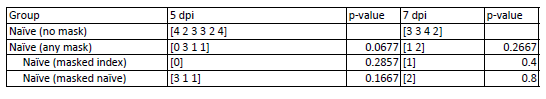

**Table S3.** Clinical scores of hamsters in the three experiments performed, with only infected hamsters included in the shown data. P-values show comparisons against the no mask experiment on the corresponding day post inoculation of the index hamster.

VI. *Analysis of weight loss data in contact animals from Ganti et al. (2022) PLoS Pathogens*

To assess support for the variolation hypothesis using one of the experimental challenge studies detailed in Ganti et al.^12^, we used patterns of weight loss in the contact animals. For every contact animal (regardless of infection status), we calculated its minimum weight over the course of the 7 days that its weight was measured (see Supplementary Figure 4 in ^12^ for the plotted data). Table S4 lists these minimum weights.

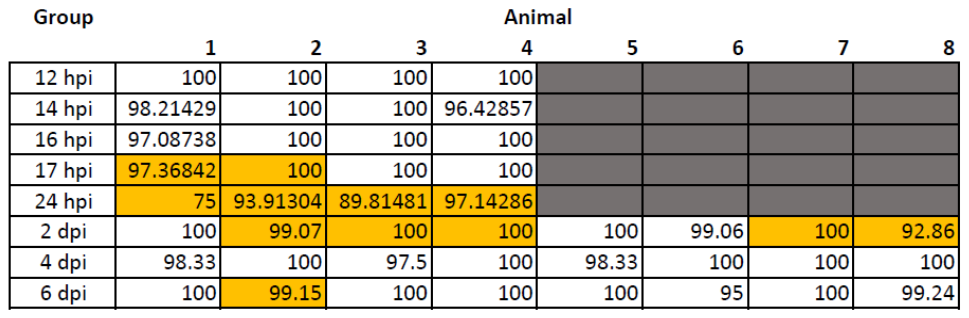

**Table S4.** Minimum weights of contact animals from the experimental study focusing on infection occurring at different time points post-inoculation of the index animal. Orange shading denotes contact animals that got infected.

We performed two-sample t-tests to determine whether contact animals that became infected differed in their minimum weight depending on the time block during which they became exposed. There were no significant differences in weight loss between infected contact animals across any pair of time blocks.

We further determined whether weight loss patterns between uninfected and infected contact animals differed from one another. Using a two-sample t-test, we found that the minimum weights of infected contacts were statistically different from, and lower, than those of uninfected contacts (*p*=0.004).

VII. *Further analysis of Dabisch et al. (2021) PLoS Pathogens*

In Figure 4A, we fit a logistic model to all individual-level data points, reproducing the dose-seroconversion relationship presented in Dabisch et al.^13^. In Figure 4B, we first fit a logistic regression model to all individual-level data points, reproducing the dose-fever development relationship presented in Dabisch et al.^13^. We then fit a logistic regression model to the subset of animals that seroconverted. This fit did not significantly alter the shape of the relationship between dose and fever development. Finally, we fit an alternative logistic regression model to the subset of animals that seroconverted. This alternative model was of the form:

Probability of fever development = *p* + (1-*p*)*r*(x), where *r*(*x*) is the standard logistic model given by

$$r\left( x \right)=\frac{1}{1+e^{-(x-\mu)/s}}$$

This alternative model allows for a non-zero probability of fever development at low inoculum doses, conditional on infection. We simultaneously estimated *p* and the two parameters of the logistic model (*μ* and *s*), with maximum likelihood estimates being *p* = 0.17, *μ* = 2.6687, and *s* = 0.0079. The Akaike Information Criterion (AIC) of the standard logistic model was 12.66. The AIC of the alternative model (which penalizes for the additional parameter) was 11.41, indicating that the alternative logistic model is statistically preferred over the standard logistic model.

Supplemental Material References

1	Lloyd-Smith JO, Schreiber SJ, Kopp PE, Getz WM. Superspreading and the effect of individual variation on disease emergence. *Nature* 2005; **438**: 355–9.

2	Ke R, Zitzmann C, Ho DD, Ribeiro RM, Perelson AS. In vivo kinetics of SARS-CoV-2 infection and its relationship with a person’s infectiousness. *Proc Natl Acad Sci USA* 2021; **118**: e2111477118.

3	Endo A, Centre for the Mathematical Modelling of Infectious Diseases COVID-19 Working Group, Abbott S, Kucharski AJ, Funk S. Estimating the overdispersion in COVID-19 transmission using outbreak sizes outside China. *Wellcome Open Res* 2020; **5**: 67.

4	Pawelek KA, Huynh GT, Quinlivan M, Cullinane A, Rong L, Perelson AS. Modeling Within-Host Dynamics of Influenza Virus Infection Including Immune Responses. *PLoS Comput Biol* 2012; **8**: e1002588.

5	Baral S, Antia R, Dixit NM. A dynamical motif comprising the interactions between antigens and CD8 T cells may underlie the outcomes of viral infections. *Proc Natl Acad Sci U S A* 2019; **116**: 17393–8.

6	Canini L, Carrat F. Population modeling of influenza A/H1N1 virus kinetics and symptom dynamics. *J Virol* 2011; **85**: 2764–70.

7	Chatterjee B, Sandhu HS, Dixit NM. The relative strength and timing of innate immune and CD8 T-cell responses underlie the heterogeneous outcomes of SARS-CoV-2 infection. Infectious Diseases (except HIV/AIDS), 2021 DOI:10.1101/2021.06.15.21258935.

8	Martin MA, Koelle K. Comment on “Genomic epidemiology of superspreading events in Austria reveals mutational dynamics and transmission properties of SARS-CoV-2”. *Sci Transl Med* 2021; **13**: eabh1803.

9	Lythgoe KA, Hall M, Ferretti L, *et al.* SARS-CoV-2 within-host diversity and transmission. *Science* 2021; **372**: eabg0821.

10	Braun KM, Moreno GK, Halfmann PJ, *et al.* Transmission of SARS-CoV-2 in domestic cats imposes a narrow bottleneck. *PLoS Pathog* 2021; **17**: e1009373.

11	Braun K, Moreno G, Wagner C, *et al.* Limited within-host diversity and tight transmission bottlenecks limit SARS-CoV-2 evolution in acutely infected individuals. Evolutionary Biology, 2021 DOI:10.1101/2021.04.30.440988.

12	Ganti K, Ferreri LM, Lee C-Y, *et al.* Timing of exposure is critical in a highly sensitive model of SARS-CoV-2 transmission. *PLoS Pathog* 2022; **18**: e1010181.

13	Dabisch PA, Biryukov J, Beck K, *et al.* Seroconversion and fever are dose-dependent in a nonhuman primate model of inhalational COVID-19. *PLoS Pathog* 2021; **17**: e1009865.
